# Service Availability and Readiness for Basic Emergency Obstetric and Newborn Care: Analysis from Nepal Health Facility Survey 2021

**DOI:** 10.1101/2023.02.15.23285987

**Authors:** Achyut Raj Pandey, Bikram Adhikari, Bipul Lamichhane, Deepak Joshi, Shophika Regmi, Bibek Kumar Lal, Sagar Dahal, Sushil Chandra Baral

## Abstract

**Background:** Although there has been significant improvement in maternal and newborn health, Nepal needs further acceleration in improving access to and coverage of services to achieve Sustainable Development Goals by 2030. In this context, we aimed to analyze availability and readiness for Basic Emergency Obstetric and Neonatal Care (BEmONC) services in Nepal using data from Nepal Health Facility Survey (NHFS), 2021.

**Methods:** We analyzed nationally representative NHFS, 2021 data to determine availability and readiness of HFs for BEmONC services based on the “Service Availability and Readiness” manual of World Health Organizations. We performed weighted descriptive and inferential analysis to account complex survey design of NHFS 2021. We summarized continuous variables with mean, standard deviation, median and interquartile range whereas categorical variables with percent and 95% confidence interval (CI). We applied simple, and multiple linear regression to determine factors associated with readiness of HFs for BEmONC services.

**Results:** The proportion of facilities with all BEmONC signal function available was 2.49% at the national level with Madhesh having the highest (5.22%) while Gandaki having the lowest (1.22%) availability. BEmONC service availability was relatively lower in rural setting and peripheral facilities. The overall readiness score for BEmONC services is 54.14, with domains of guidelines/staff, equipment/supplies, and medicines/commodities having scores of 21.16, 76.76, and 64.50 respectively. In multivariable analysis, level of facilities, province, urban rural setting, ecological belt, presence of external supervision, 24-hour duty schedule and number of beds were found to have statistically significant association with BEmONC service readiness.

**Conclusion:** The readiness of HFs for BEmONC services in Nepal is relatively poor and can be improved by increasing service provision, expanding service hours to 24-hours a day, increasing availability of essential medicines and equipment, enhanced monitoring, and conducting periodic review of maternal and newborn deaths at HFs.

## Background

Sustainable Development Goal (SDG) 3.1 aims to reduce maternal mortality to fewer than 70 per 100,000 live births globally [1]. Nepal has made remarkable progress in maternal health over the previous few decades. Between 1990 and 2017, the average yearly rate of change in the maternal mortality ratio (MMR) was −2.9% [2]. However, the rate of reduction has been rather gradual in recent years. Since 2005, Nepal’s MMR has decreased by 0.9%. With the current rate of decline, MMR in Nepal is anticipated to reach 199.2 by 2030 [2] which is almost three times higher than the target of 70 set by the 2030 SDG.

Timely access to emergency obstetric and newborn care (EmONC) services is one of the key drivers to reduce maternal and neonatal mortality globally [3,4]. To get on the right track in decreasing preventable maternal mortality and to help countries in tracking progress, the World Health Organization (WHO) and UNFPA announced five essential coverage objectives and milestones in 2020, of which under “Every Pregnant Woman with obstetric complications” heading, global target was to ensure at least 60% of the population physically access the nearest emergency obstetric and newborn health facility within two hours of travel time and 80% of countries are targeted to have at least 50% of their population able to access the nearest EmONC services physically within 2 hours of travel time [3]. Furthermore, it is universally recommended to treat all cases of serious direct obstetric difficulties in emergency obstetric care facilities having a case fatality rate of less than 1% among women treated in emergency obstetric care facilities[5].

In context of Nepal, the Safe Motherhood and Newborn Health (SMNH) Road Map 2030, recommends encouraging all women to give birth at a BEmONC/CEmONC facility if such facilities are within 2 hours walking distance from their house. The strategy suggests that existing BEmONC sites should be fully operationalized, with selected birthing facilities being upgraded to BEmONC sites [6]. In line with the global recommendations, SMNH roadmap also recommends that the met need of EOC should be 100% [6]. However, the latest routine data from Department of Health Services reveal that the nationwide availability of EOC met need hovered around 11-12% for three fiscal years from 2017/18 to 2019/20, then dropped to roughly 8% in 2020/21. These figures on met need of EOC are concerning and signals reduction in utilization or delivery of complication treatment during pregnancy[7]. Data show variation across the seven provinces with met need of EOC ranging from 3.5% to 10.5% with highest in Lumbini province (10.5%) and lowest in Madhesh (3.5%) [7].

A study by Bhutta et al in 2014 showed that around 150000 maternal deaths, 550,000 stillbirths and 790,000 neonatal deaths could be prevented globally by scaling up the care during labour and childbirths including complications.[8] Similarly, Studies had estimated that approximately 15-20% of all pregnancies are complicated and require hospital attention [9,10] thus requiring health facilities providing delivery services to deal with the complicated cases in-order to handle such case as they arise. According to a previous systematic review and delphi estimation, skilled childbirth care alone would prevent 25% of intrapartum-related neonatal deaths compared to no skilled care.[11] Similarly, Basic and comprehensive emergency obstetric care was estimated to prevent 40% and 85%, respectively, neonatal deaths due to intrapartum events.[8,11] A rigorous assessment of the extent to which facilities offering delivery services are prepared to provide emergency obstetric care becomes crucial. In this context, we have attempted to analyze the service availability and readiness in Nepalese context using data from Nepal Health Facility Survey (NHFS) 2021 [12].

## Methods

We performed secondary data analysis of a nationally representative data from NHFS 2021 to determine service availability and readiness of EmONC among facilities offering normal vaginal delivery services. [12] The NHFS 2021 was implemented by New ERA, a research firm, with the support from MOHP and ICF International.

### Sample and sampling techniques

The NHFS 2021 collected data from a sample of facilities managed by the government and private not-for-profit non-governmental organizations (NGOs), private for-profit organizations, and mission/faith organizations in all 77 districts of the country with the objective of providing a comprehensive picture of the strengths and weaknesses of the service delivery environment. From the main list of 7,598 health facilities categorized according to management authority (i.e., governmental or non-governmental), a total of 1,917 health facilities were excluded using predefined exclusion criteria (polyclinics or hospitals that provided stand-alone specialist services such as cancer and heart treatment). Hospitals, PHCCs, HPs, CHUs, stand-alone HTCs, and UHCs were divided into six groups. During the selection process, several sample procedures were used on various types of facilities. In final stage 1,633 health facilities chosen at random were sampled by NHFS 2021 using equal probability systematic sampling with sample allocation [12]. The sampling approach is described in depth elsewhere [12]. From the set of 1633 facilities, this analysis has been carried out among 786 facilities offering normal vaginal delivery.

### Data collection

The data collection of NHFS 2021 was performed between 27 January 2021, and 28 September, 2021, which was halted for three months from May to July due to COVID-19-imposed lockdowns in Nepal [12]. The Nepal Health Facility Survey 2021 used four main tools for data collection, namely a) facility inventory questionnaire, b) healthcare worker interview questionnaire, c) observational protocol (for ANC, family planning, sick child services, labor, and childbirth) and d) exit interview questionnaires (for ANC and family planning clients and for caretakers of sick children whose consultations were observed. For this article, we analyzed variables from the Facility Inventory Questionnaire and one variable on staff training from the Health Provider Questionnaire.

### Outcome variables and measurement

The variables for services availability and readiness of health facilities for BEmONC were selected based on the WHO service availability and readiness (SARA) manual [13].

### Services availability

The service availability of health facilities for BEmONC was measured based on seven signal functions of delivery and newborn care services which included parenteral antibiotics, parenteral oxytocin, parenteral anticonvulsant, assisted vaginal delivery, manual removal of placenta, removal of retained product of conception, and neonatal resuscitation. The seven signal functions measure the capacity of health facilities to treat obstetric and newborn emergencies[3]. The health facility was defined to have availability of functional signal function of delivery and newborn care if the service was available and provided within past 3 months preceding the survey.

### Services Readiness

The service readiness of health facilities for BEmONC was measured based on the availability and functioning of items categorized into three domains-staff and guidelines (2 items), essential equipment and supplies (14 items) and essential medicine and commodities (11 items). The list of tracer items of each domain is present in the **<u>supplementary file 1</u>**. The readiness score of health facilities to provide BEOmNC services was calculated using Service Availability and Readiness Assessment manual of the World Health Organization [13]. The items in each domain were recoded as binary variable, taking 1 for availability and 0 for absence of the item in the facility. The mean score for each domain was computed by adding items that are divided by number of items and multiplied by 100. The average of the score from three domains was the readiness score.

### Independent variable

The independent variables included ecological region (hill / mountain / terai), location of facility (rural / urban), province (Province 1 / Madhesh / Bagmati / Gandaki /Lumbini / Karnali / Sudurpaschim), type of facility (federal, provincial hospital / peripheral facilities / private hospital), presence of external supervision in past four months (present / absent), duty schedule or call list for 24-hour staff assignment (yes / no), reviews of maternal or newborn deaths (reviewed / not reviewed), system of determining and reviewing clients’ opinion (reviewed / not reviewed), quality assurance activities (performed /not performed), frequency of health facility administrative and management meeting (none / sometimes / monthly), number of delivery beds and number of total health workers.

The rural-urban classification of settings was based on the municipality types in which the health facilities are situated. Facilities located in rural municipality were classified as ‘rural’ whereas facilities located in municipality, sub-metropolitan and metropolitan city were classified as ‘urban’. The hospitals under federal or provincial government were classified as federal or provincial hospitals, facilities under local governments which include local hospital, primary health care center and health posts were classified as peripheral facilities, and the hospitals owned by private sectors were classified as private hospitals. Facilities that received any kind of monitoring or supervision from federal, provincial, or municipal authorities in the past four months were considered to have external supervision. The facilities are said to have duty schedule or call list for 24-hour staff assignment if enumerators observed duty schedule or call list for 24-hour staff coverage The facilities are said to have to have reviewed of maternal or newborn deaths if those facilities regularly reviewed maternal and new-born deaths. The facilities that had a system for determining client opinions, procedures for reviewing client opinions, and reports of recent reviews of client opinions were considered to have performed a review of client opinions. Health facilities that routinely carry out quality assurance activities and have documentation of recent quality assurance activities, such as quality assurance reports, supervisory checklists, mortality reviews, or audits of records, were considered to have performed quality assurance activities. Health facilities that reported “no” for routine management/administrative meetings were classified as “none”, while those that stated, “monthly or more often” were classified as “monthly or more often” and those that reported “irregular or every 2-6 months” were classified as “sometimes”.

### Statistical analysis

We used R version 4.2.0[14] for the statistical analysis and performed weighted analysis using “survey” {Lumley, 2012 #21} package to account complex survey design of 2021 NHFS. Continuous variables were summarized with mean, standard deviation (SD), median and interquartile range (IQR) whereas categorical variables were summarized with weighted percent and 95 percent confidence interval (CI). Univariate and multivariate weighted linear regression analysis were used to determine association of readiness of health facilities to BEmONC with independent variables ecological region, location of facility, province, type of facility, presence of external supervision in past four months, duty schedule or call list for 24-hour staff assignment, reviews of maternal or newborn deaths, system of determining and reviewing clients’ opinion, quality assurance activities, frequency of health facility meeting, number of delivery beds and number of total health workforce. We used variance inflation factor (VIF) to check for multi-collinearity among independent variables. A p-value of less than 0.05 was considered statistically significant.

### Ethical approval

The study involves secondary analysis of data from NHFS 2021; hence no ethical approval was sought for this study. In the original survey NHFS 2021, ethical clearance was obtained from the Ethical Review Board of Nepal Health Research Council and ICF International and the details of the methodology is published in the survey report [12].

## Results

Table 1 presents the characteristics of health facilities offering normal vaginal delivery. Of the total facilities offering normal vaginal delivery, 61.36% facilities were from Hilly region, 16.96% facilities were from mountain region and 21.68% facilities were from Terai region. In terms of province, 16.67% of facilities were from Province 1, 7.62% from Madhesh, 18.84% from Bagmati, 11.40% from Gandaki, 16.90% from Lumbini, 12.42% from Karnali and 16.14% from Sudurpaschim Province. Based on ownership, 92.4% facilities were public while the remaining 7.6% facilities were private. Approximately 95% (94.84%) facilities were peripheral facilities whereas 5.16% facilities were hospitals and less than one quarter (21.81%) of the facilities had 24-hour duty schedule. Furthermore, external supervision was present in 68.13% facilities, quality assurance activities were performed in 31% facilities, review of maternal and newborn deaths was done in 32.67% facilities and client opinion was reviewed in 5.34% facilities. Among the facilities, Special Newborn Care Unit (SNCUs) were present in 5.96% facilities and on an average 27 health workers and 1.30 beds were present per facility.

**Table 1:**
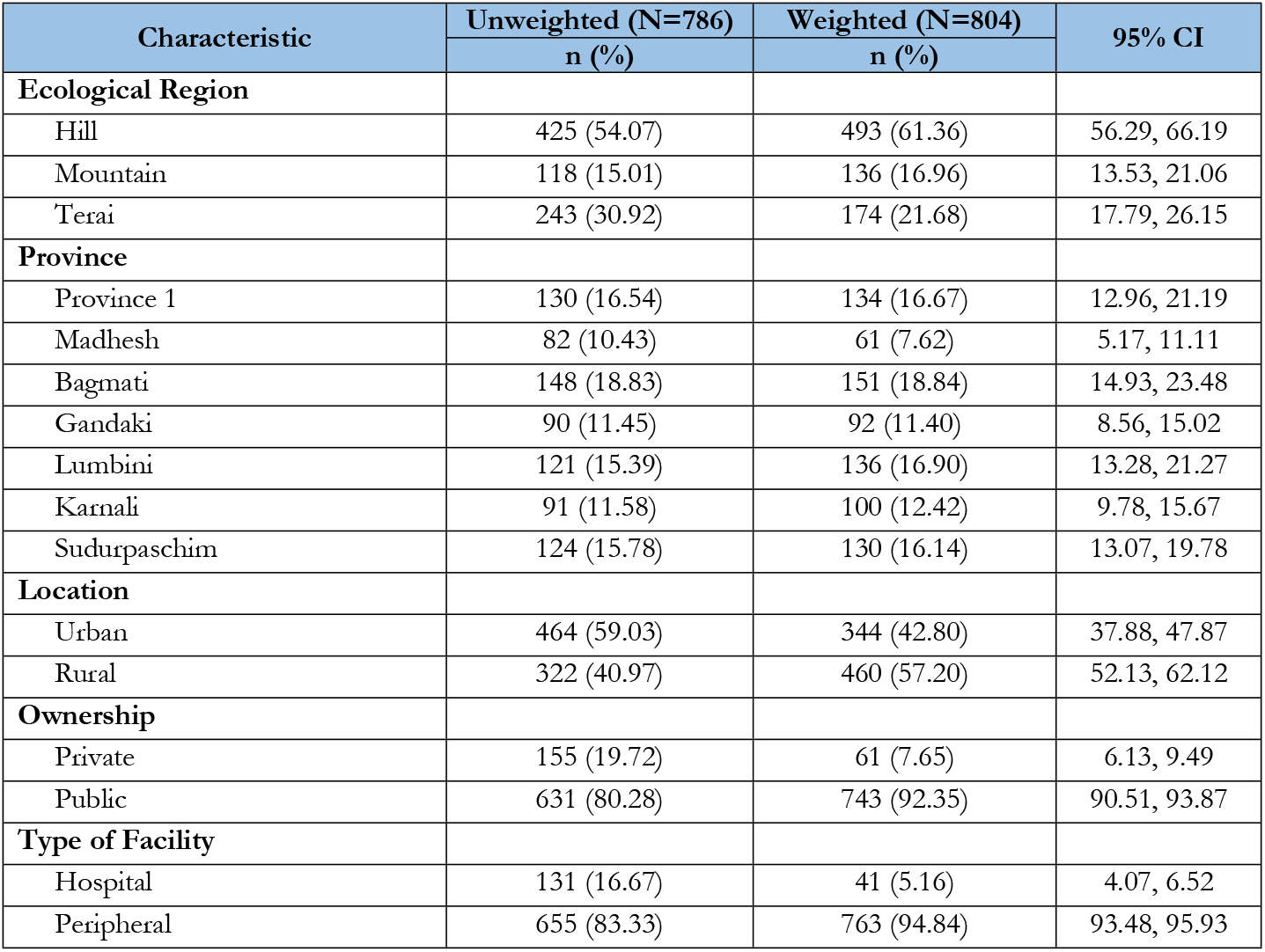
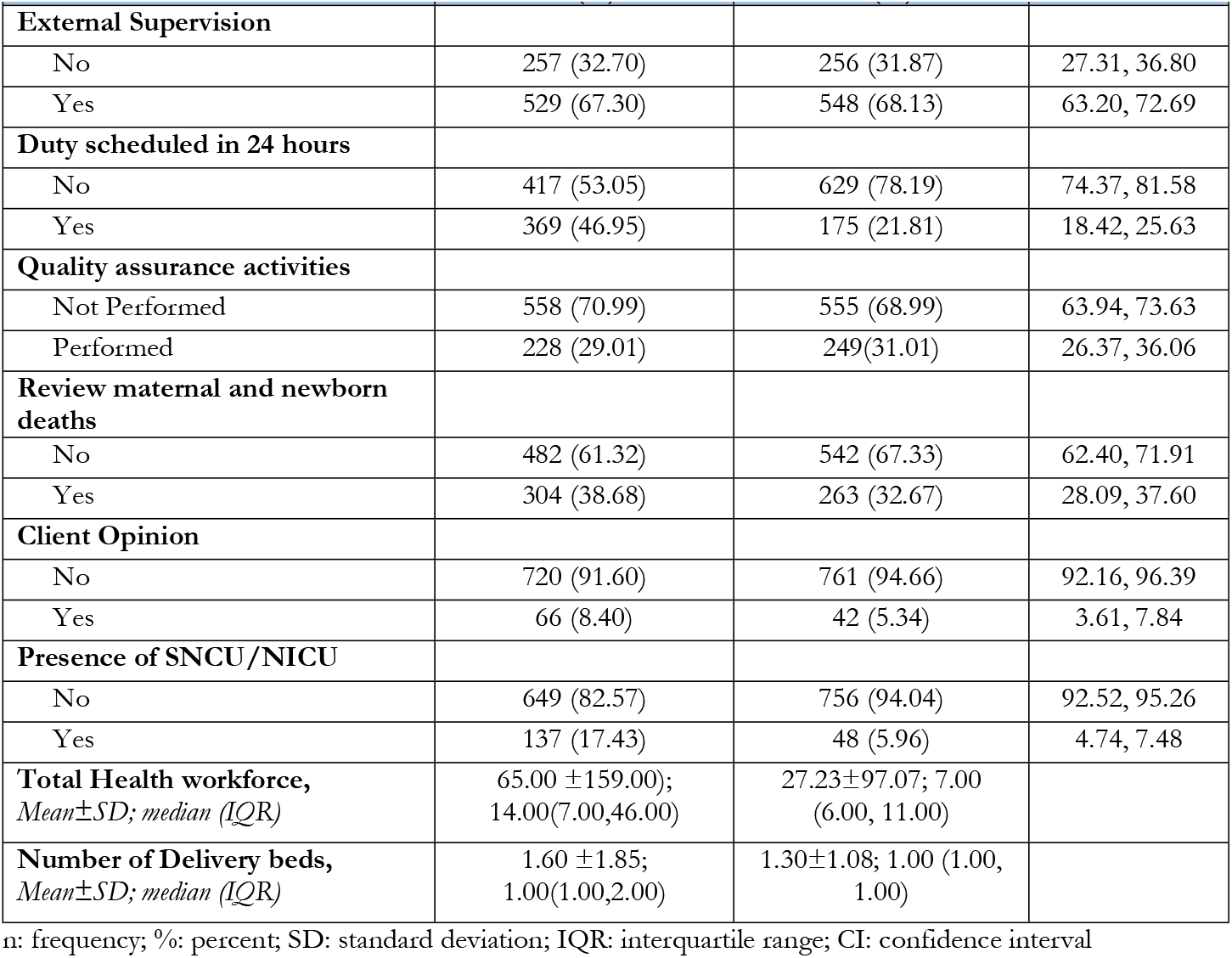
Distribution of facilities offering normal vaginal delivery.

| Characteristic | Unweighted (N=786) | Weighted (N=804) | 95% CI |
| --- | --- | --- | --- |
|  | n (%) | n (%) |  |
| **Ecological Region** |  |  |  |
| Hill | 425 (54.07) | 493 (61.36) | 56.29, 66.19 |
| Mountain | 118 (15.01) | 136 (16.96) | 13.53, 21.06 |
| Terai | 243 (30.92) | 174 (21.68) | 17.79, 26.15 |
| **Province** |  |  |  |
| Province 1 | 130 (16.54) | 134 (16.67) | 12.96, 21.19 |
| Madhesh | 82 (10.43) | 61 (7.62) | 5.17, 11.11 |
| Bagmati | 148 (18.83) | 151 (18.84) | 14.93, 23.48 |
| Gandaki | 90 (11.45) | 92 (11.40) | 8.56, 15.02 |
| Lumbini | 121 (15.39) | 136 (16.90) | 13.28, 21.27 |
| Karnali | 91 (11.58) | 100 (12.42) | 9.78, 15.67 |
| Sudurpaschim | 124 (15.78) | 130 (16.14) | 13.07, 19.78 |
| **Location** |  |  |  |
| Urban | 464 (59.03) | 344 (42.80) | 37.88, 47.87 |
| Rural | 322 (40.97) | 460 (57.20) | 52.13, 62.12 |
| **Ownership** |  |  |  |
| Private | 155 (19.72) | 61 (7.65) | 6.13, 9.49 |
| Public | 631 (80.28) | 743 (92.35) | 90.51, 93.87 |
| **Type of Facility** |  |  |  |
| Hospital | 131 (16.67) | 41 (5.16) | 4.07, 6.52 |
| Peripheral | 655 (83.33) | 763 (94.84) | 93.48, 95.93 |
|  | n (%) | n (%) |  |
| **External Supervision** |  |  |  |
| No | 257 (32.70) | 256 (31.87) | 27.31, 36.80 |
| Yes | 529 (67.30) | 548 (68.13) | 63.20, 72.69 |
| **Duty scheduled in 24 hours** |  |  |  |
| No | 417 (53.05) | 629 (78.19) | 74.37, 81.58 |
| Yes | 369 (46.95) | 175 (21.81) | 18.42, 25.63 |
| **Quality assurance activities** |  |  |  |
| Not Performed | 558 (70.99) | 555 (68.99) | 63.94, 73.63 |
| Performed | 228 (29.01) | 249(31.01) | 26.37, 36.06 |
| **Review maternal and newborn deaths** |  |  |  |
| No | 482 (61.32) | 542 (67.33) | 62.40, 71.91 |
| Yes | 304 (38.68) | 263 (32.67) | 28.09, 37.60 |
| **Client Opinion** |  |  |  |
| No | 720 (91.60) | 761 (94.66) | 92.16, 96.39 |
| Yes | 66 (8.40) | 42 (5.34) | 3.61, 7.84 |
| **Presence of SNCU/NICU** |  |  |  |
| No | 649 (82.57) | 756 (94.04) | 92.52, 95.26 |
| Yes | 137 (17.43) | 48 (5.96) | 4.74, 7.48 |
| **Total Health workforce,**<br>**Mean±SD; median (IQR)** | 65.00 ±159.00;<br>14.00(7.00,46.00) | 27.23±97.07; 7.00<br>(6.00, 11.00) |  |
| **Number of Delivery beds,**<br>**Mean±SD; median (IQR)** | 1.60 ±1.85;<br>1.00(1.00,2.00) | 1.30±1.08; 1.00 (1.00,<br>1.00) |  |

n: frequency; %: percent; SD: standard deviation; IQR: interquartile range; CI: confidence interval

**Table 2:**
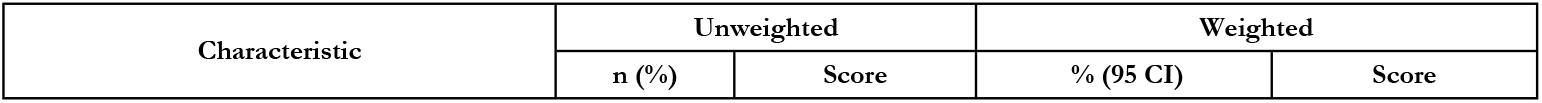
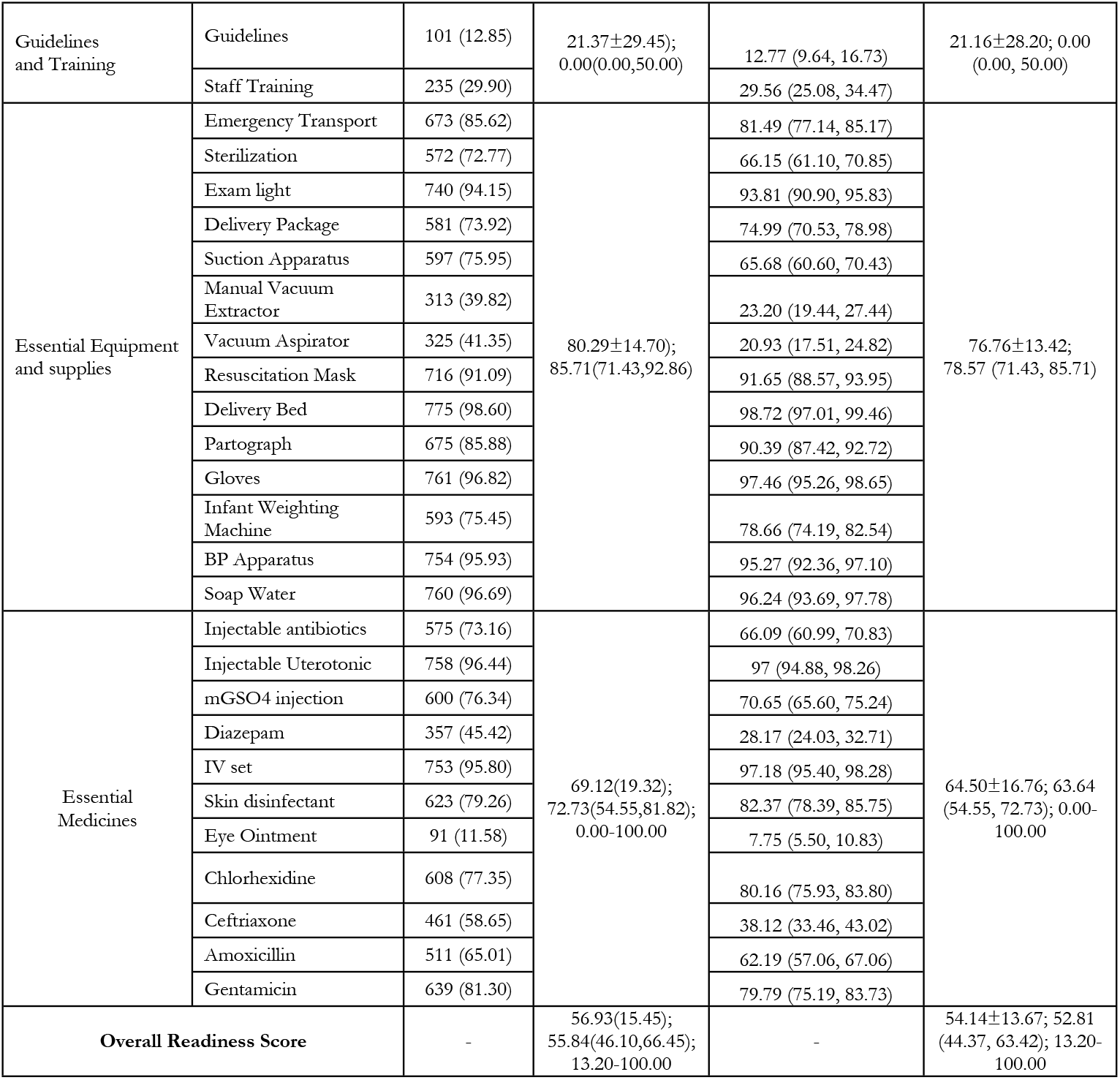
Readiness for EMONC service

| Characteristic | Unweighted |  | Weighted |  |
| --- | --- | --- | --- | --- |
|  | n (%) | Score | % (95 CI) | Score |

|  |  |  |  |  |  |
| --- | --- | --- | --- | --- | --- |
| Guidelines and Training | Guidelines | 101 (12.85) | 21.37±29.45);<br>0.00(0.00,50.00) | 12.77 (9.64, 16.73) | 21.16±28.20; 0.00<br>(0.00, 50.00) |
|  | Staff Training | 235 (29.90) |  | 29.56 (25.08, 34.47) |  |
| Essential Equipment and supplies | Emergency Transport | 673 (85.62) | 80.29±14.70);<br>85.71(71.43,92.86) | 81.49 (77.14, 85.17) | 76.76±13.42;<br>78.57 (71.43, 85.71) |
|  | Sterilization | 572 (72.77) |  | 66.15 (61.10, 70.85) |  |
|  | Exam light | 740 (94.15) |  | 93.81 (90.90, 95.83) |  |
|  | Delivery Package | 581 (73.92) |  | 74.99 (70.53, 78.98) |  |
|  | Suction Apparatus | 597 (75.95) |  | 65.68 (60.60, 70.43) |  |
|  | Manual Vacuum Extractor | 313 (39.82) |  | 23.20 (19.44, 27.44) |  |
|  | Vacuum Aspirator | 325 (41.35) |  | 20.93 (17.51, 24.82) |  |
|  | Resuscitation Mask | 716 (91.09) |  | 91.65 (88.57, 93.95) |  |
|  | Delivery Bed | 775 (98.60) |  | 98.72 (97.01, 99.46) |  |
|  | Partograph | 675 (85.88) |  | 90.39 (87.42, 92.72) |  |
|  | Gloves | 761 (96.82) |  | 97.46 (95.26, 98.65) |  |
|  | Infant Weighting Machine | 593 (75.45) |  | 78.66 (74.19, 82.54) |  |
|  | BP Apparatus | 754 (95.93) |  | 95.27 (92.36, 97.10) |  |
|  | Soap Water | 760 (96.69) |  | 96.24 (93.69, 97.78) |  |
| Essential Medicines | Injectable antibiotics | 575 (73.16) | 69.12(19.32);<br>72.73(54.55,81.82);<br>0.00-100.00 | 66.09 (60.99, 70.83) | 64.50±16.76; 63.64<br>(54.55, 72.73); 0.00-100.00 |
|  | Injectable Uterotonic | 758 (96.44) |  | 97 (94.88, 98.26) |  |
|  | mGSO4 injection | 600 (76.34) |  | 70.65 (65.60, 75.24) |  |
|  | Diazepam | 357 (45.42) |  | 28.17 (24.03, 32.71) |  |
|  | IV set | 753 (95.80) |  | 97.18 (95.40, 98.28) |  |
|  | Skin disinfectant | 623 (79.26) |  | 82.37 (78.39, 85.75) |  |
|  | Eye Ointment | 91 (11.58) |  | 7.75 (5.50, 10.83) |  |
|  | Chlorhexidine | 608 (77.35) |  | 80.16 (75.93, 83.80) |  |
|  | Ceftriaxone | 461 (58.65) |  | 38.12 (33.46, 43.02) |  |
|  | Amoxicillin | 511 (65.01) |  | 62.19 (57.06, 67.06) |  |
|  | Gentamicin | 639 (81.30) |  | 79.79 (75.19, 83.73) |  |
| Overall Readiness Score |  | - | 56.93(15.45);<br>55.84(46.10,66.45);<br>13.20-100.00 | - | 54.14±13.67; 52.81<br>(44.37, 63.42); 13.20-100.00 |

**Table 3:**
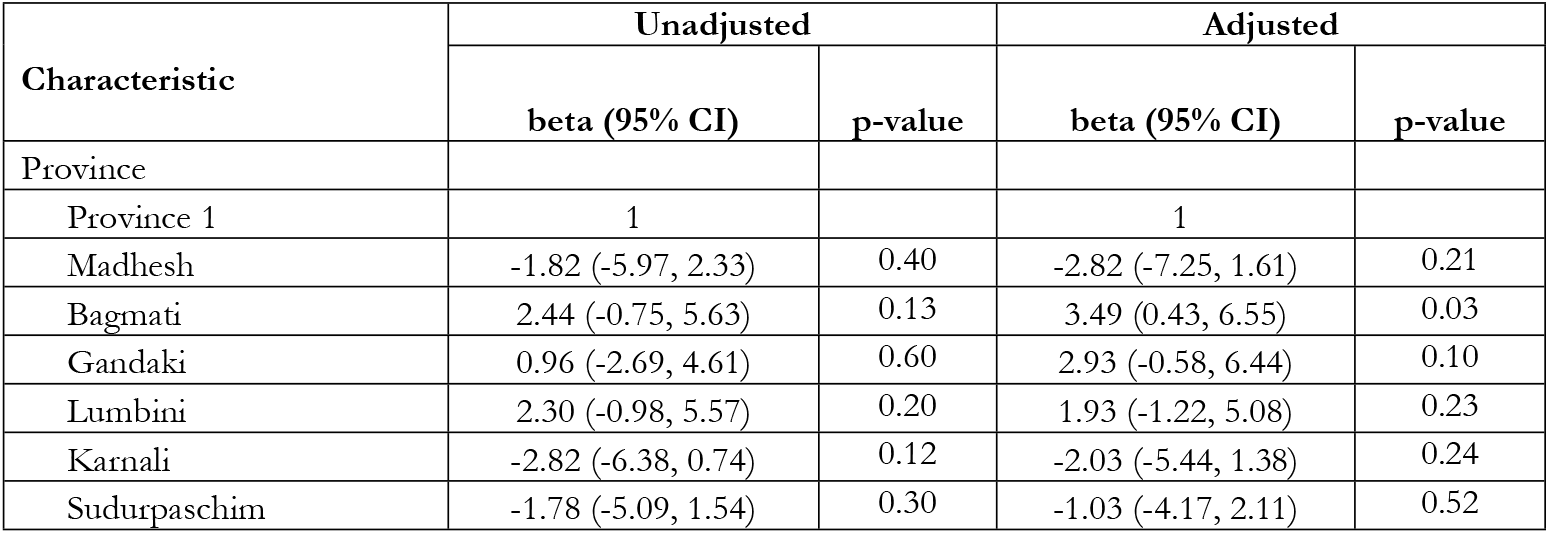
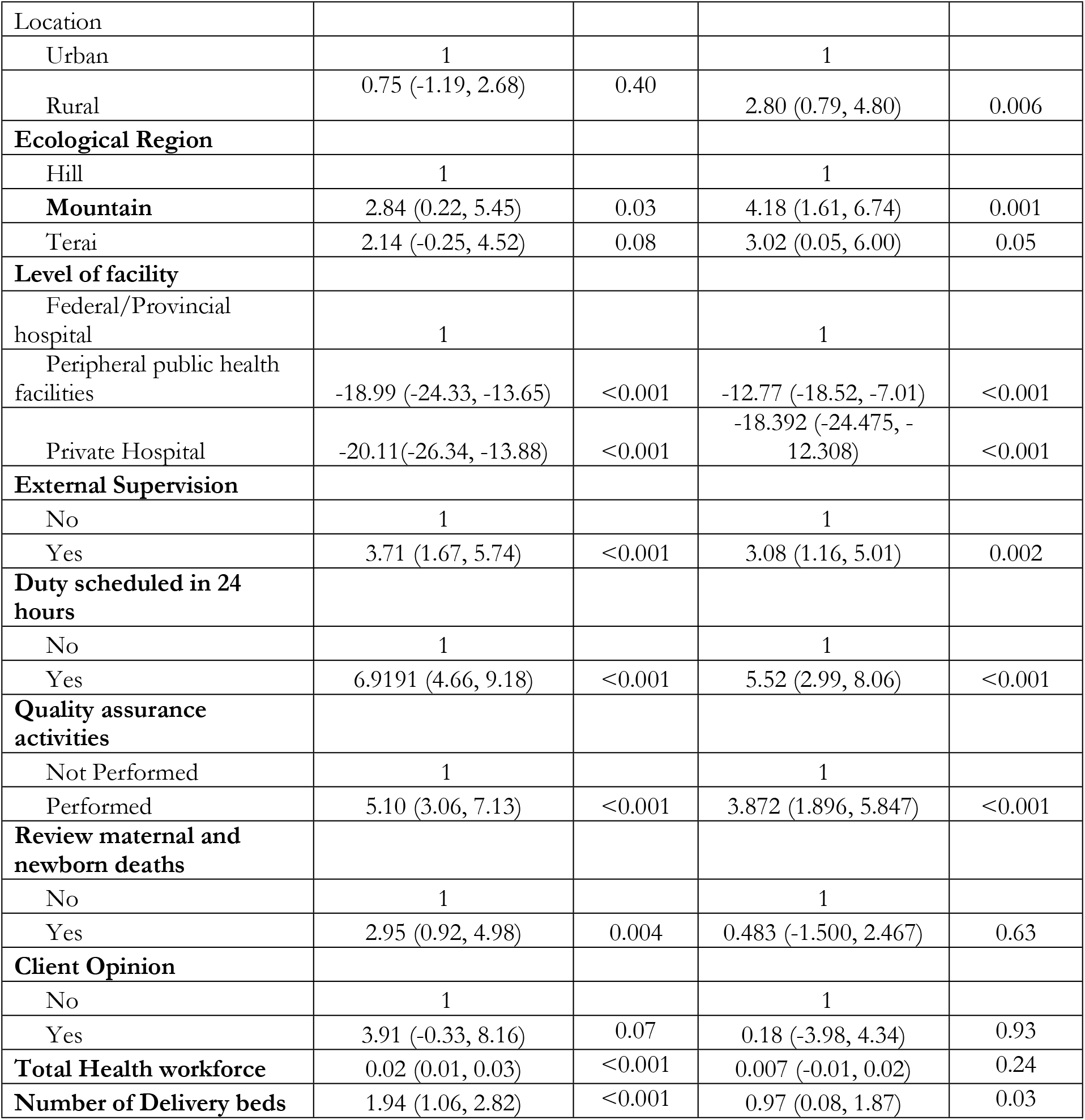
Factors associated with BEmONC service readiness.

| Characteristic | Unadjusted |  | Adjusted |  |
| --- | --- | --- | --- | --- |
|  | beta (95% CI) | p-value | beta (95% CI) | p-value |
| Province |  |  |  |  |
| Province 1 | 1 |  | 1 |  |
| Madhesh | -1.82 (-5.97, 2.33) | 0.40 | -2.82 (-7.25, 1.61) | 0.21 |
| Bagmati | 2.44 (-0.75, 5.63) | 0.13 | 3.49 (0.43, 6.55) | 0.03 |
| Gandaki | 0.96 (-2.69, 4.61) | 0.60 | 2.93 (-0.58, 6.44) | 0.10 |
| Lumbini | 2.30 (-0.98, 5.57) | 0.20 | 1.93 (-1.22, 5.08) | 0.23 |
| Karnali | -2.82 (-6.38, 0.74) | 0.12 | -2.03 (-5.44, 1.38) | 0.24 |
| Sudurpaschim | -1.78 (-5.09, 1.54) | 0.30 | -1.03 (-4.17, 2.11) | 0.52 |
| Location |  |  |  |  |
| Urban | 1 |  | 1 |  |
| Rural | 0.75 (-1.19, 2.68) | 0.40 | 2.80 (0.79, 4.80) | 0.006 |
| **Ecological Region** |  |  |  |  |
| Hill | 1 |  | 1 |  |
| Mountain | 2.84 (0.22, 5.45) | 0.03 | 4.18 (1.61, 6.74) | 0.001 |
| Terai | 2.14 (-0.25, 4.52) | 0.08 | 3.02 (0.05, 6.00) | 0.05 |
| **Level of facility** |  |  |  |  |
| Federal/Provincial hospital | 1 |  | 1 |  |
| Peripheral public health facilities | -18.99 (-24.33, -13.65) | <0.001 | -12.77 (-18.52, -7.01) | <0.001 |
| Private Hospital | -20.11(-26.34, -13.88) | <0.001 | -18.392 (-24.475, -12.308) | <0.001 |
| **External Supervision** |  |  |  |  |
| No | 1 |  | 1 |  |
| Yes | 3.71 (1.67, 5.74) | <0.001 | 3.08 (1.16, 5.01) | 0.002 |
| **Duty scheduled in 24 hours** |  |  |  |  |
| No | 1 |  | 1 |  |
| Yes | 6.9191 (4.66, 9.18) | <0.001 | 5.52 (2.99, 8.06) | <0.001 |
| **Quality assurance activities** |  |  |  |  |
| Not Performed | 1 |  | 1 |  |
| Performed | 5.10 (3.06, 7.13) | <0.001 | 3.872 (1.896, 5.847) | <0.001 |
| **Review maternal and newborn deaths** |  |  |  |  |
| No | 1 |  | 1 |  |
| Yes | 2.95 (0.92, 4.98) | 0.004 | 0.483 (-1.500, 2.467) | 0.63 |
| **Client Opinion** |  |  |  |  |
| No | 1 |  | 1 |  |
| Yes | 3.91 (-0.33, 8.16) | 0.07 | 0.18 (-3.98, 4.34) | 0.93 |
| **Total Health workforce** | 0.02 (0.01, 0.03) | <0.001 | 0.007 (-0.01, 0.02) | 0.24 |
| **Number of Delivery beds** | 1.94 (1.06, 2.82) | <0.001 | 0.97 (0.08, 1.87) | 0.03 |

Figure 1 presents the availability of seven signal functions among facilities offering normal vaginal delivery. Of total facilities offering normal vaginal delivery services,36.45% had parenteral antibiotics, 88.2% had parenteral oxytocin, 8.87% had parenteral anticonvulsants, 8.13% had assisted vaginal delivery, 36.72% had service for manual delivery of placenta, 26.37% had service for removal for retained placenta and 29.56% had neonatal resuscitation service.

**Figure 1:**
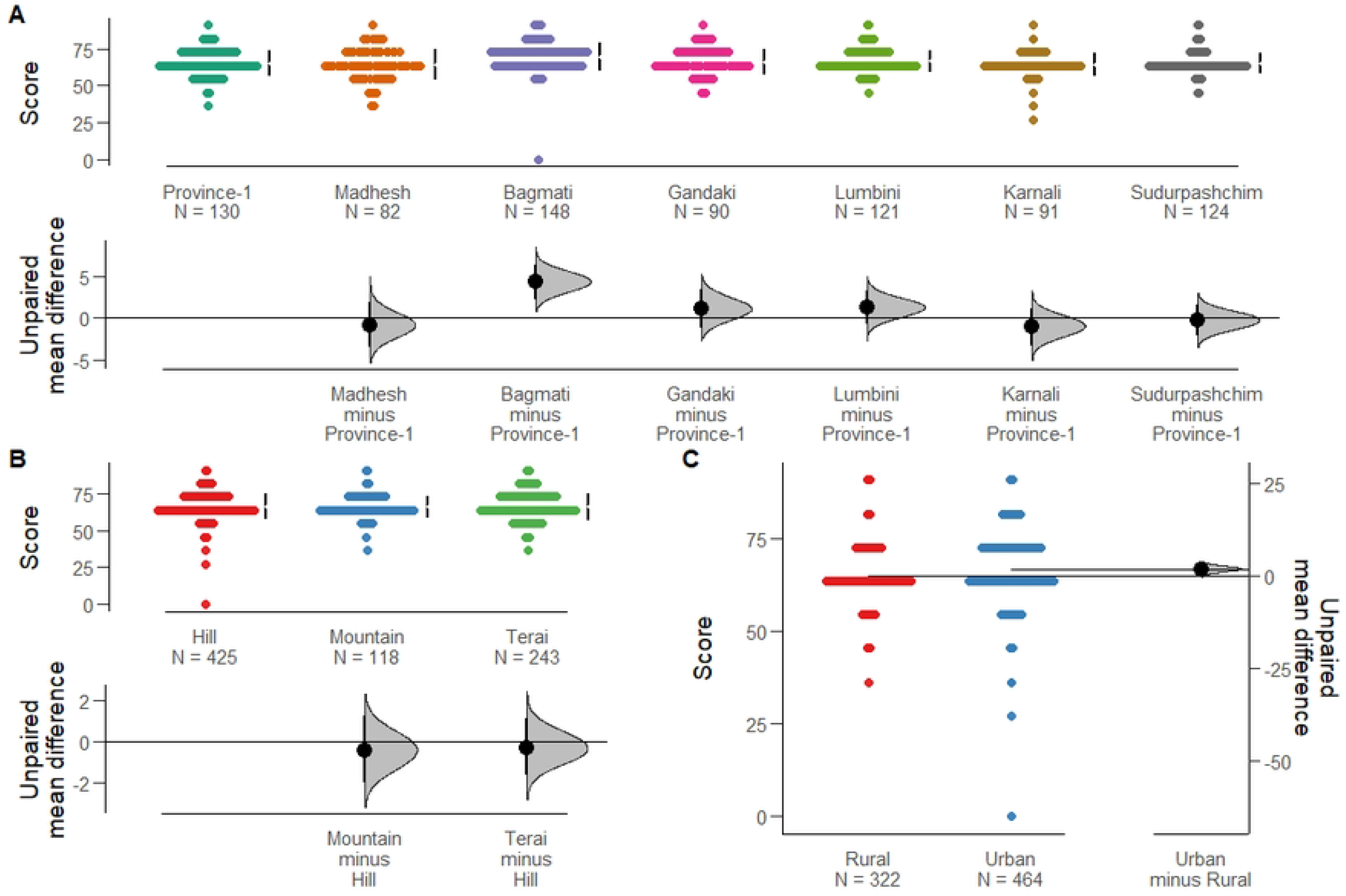
Availability of BEmONC signal functions.

Figure-2 presents distribution of facilities with availability of all seven signal functions by province, ecological region, setting, type of health facility and ownership. Around 2.49% of facilities at national level had all seven signal functions available. The availability of BEmONC signal functions varied substantially by province. For example, 5.22% of facilities in Madhesh province had all seven signal functions available while only 1.22% facilities in Gandaki, 1.51% in Sudurpaschim, 1.68% in Karnali, 2.22% in Bagmati, 2.77% in Lumbini and 3.69% in Province 1 had the seven signal functions. In terms of ecological belt, the proportion of facilities with all signal function available was 6.39% in Terai, 1.58% in hill and 0.82% in mountain region. Similarly, 5.74% of facilities in urban area and 0.06% of facilities in rural area had all seven signal functions available. Around 45.04% of provincial/federal hospitals, and 0.31% of peripheral health facilities had all seven BEmONC signal functions available. The availability of all seven signal functions was highest in public facilities accounting 1.83% in public health facilities and 10.52% in private facilities.

**Figure 2:**
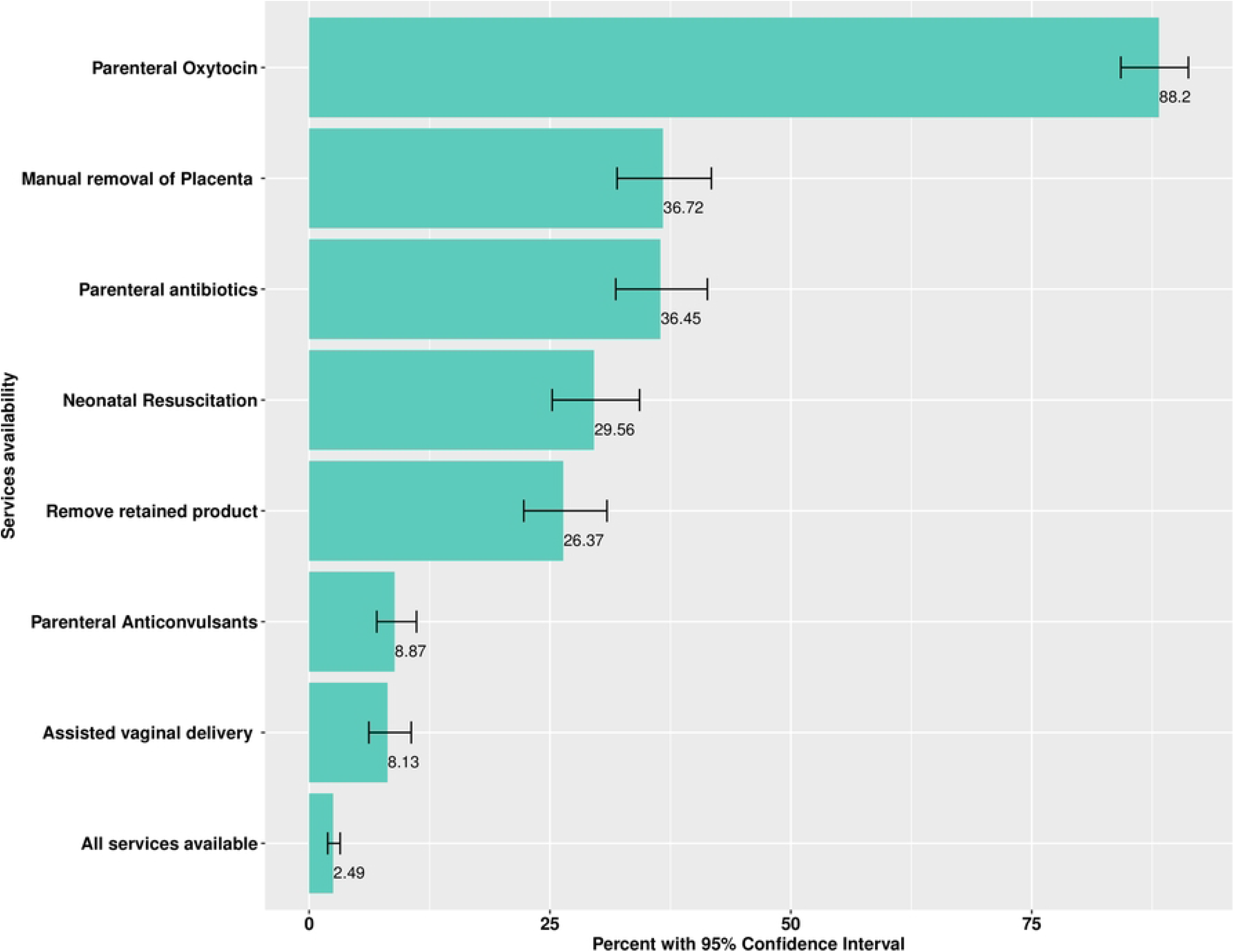
Distribution of facilities with all seven signal functions available.

Figure 3 presents the BEmONC preparedness score at aggregate level and in three domains: guidelines and staffs, equipment, essential medicines, and commodities. Each of these domains were scored in 33.33 points to obtain a national aggregate score in 100. However, for the ease in interpretation, score in each of these three domains is scaled to 100. The aggregate weighted BEmONC service readiness score was 54.14. The weighted readiness score for guideline and staffs, equipment and supplies and essential medicine and commodities were 21.16, 76.76 and 64.50 respectively.

**Figure 3:**
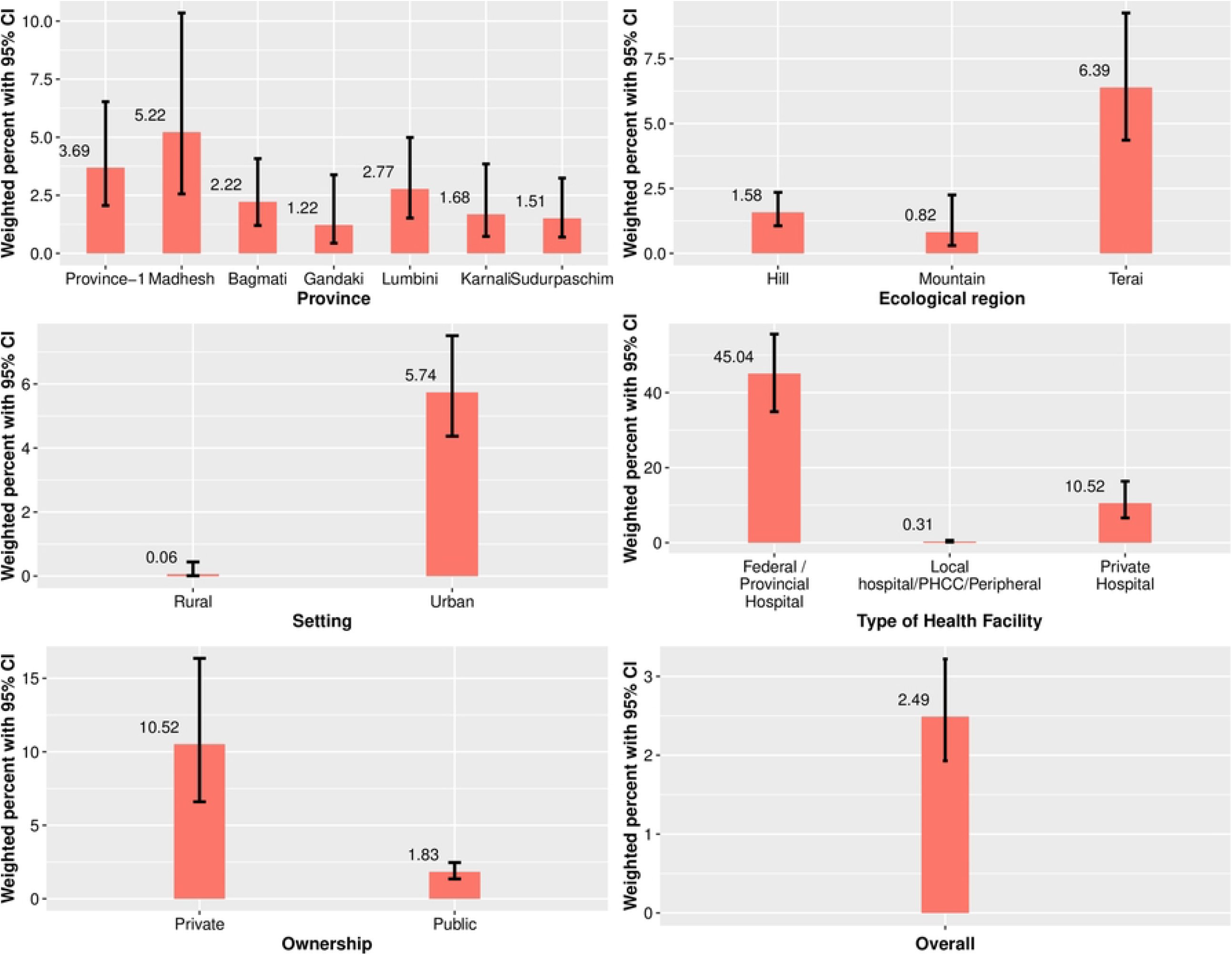
Readiness score for three domains of service readiness.

With a total score of 21.16 out of 100, guidelines and training are the core areas that need improvement. Only 12.77% of the facilities were prepared in terms of the availability of guidelines for service delivery. Around 29.56% of facilities had at least one service provider trained on delivery services (skill births attendant (SBA), advanced SBA, maternal and newborn health update, routine care during labor and normal vaginal delivery, active management of third stage labor (AMTSL)) in last 24 months preceding the survey. Similarly, the proportion of facilities having at least one dedicated bed for safe delivery was 98.72%. Furthermore, 97.46% of health facilities had gloves, 96.24% had soap water and 95.27 had BP apparatus. Nine out of ten facilities offering normal vaginal delivery had sterilization equipment examination light (93.81%), resuscitation mask (91.65%) and partograph (90.39). Similarly, two third or more of the facilities had emergency transport (81.49%), sterilization (66.15%), delivery package (74.99%), suction apparatus (65.68%) and infant weighing machine (78.66%). However, fewer than one third of the total facilities had a manual vacuum extractor (23.20%) and vacuum aspirator (20.93%).

Among essential medicines, injectable Uterotonic (97%), IV set (97.18%), skin disinfectants (82.37), Chlorhexidine (80.16%) and Gentamycin (79.79%) were the five most commonly available medicine in facilities offering normal vaginal delivery. Similarly, Injectable antibiotics were available in 66.09% facilities, ceftriaxone in 66.09% facilities and Amoxicillin in 62.19% facilities. Eye ointment was available in only 7.75 facilities and 28.17 facilities had Diazepam. (Table 4)

Figure 3 presents the BEmONC preparedness score at aggregate level and in three domains: guidelines and staffs, equipment, essential medicines, and commodities. Each of these domains were scored in 33.33 points to obtain a national aggregate score in 100. However, for the ease in interpretation, score in each of these three domains is scaled to 100. The aggregate weighted BEmONC service readiness score was 54.14. The weighted readiness score for guideline and staffs, equipment and supplies and essential medicine and commodities were 21.16, 76.76 and 64.50 respectively.

With a total score of 21.16 out of 100, guidelines and training are the core areas that need improvement. Only 12.77% of the facilities were prepared in terms of the availability of guidelines for service delivery. Around 29.56% of facilities had at least one service provider trained on delivery services (skill births attendant (SBA), advanced SBA, maternal and newborn health update, routine care during labor and normal vaginal delivery, active management of third stage labor (AMTSL)) in last 24 months preceding the survey. Similarly, the proportion of facilities having at least one dedicated bed for safe delivery was 98.72%. Furthermore, 97.46% of health facilities had gloves, 96.24% had soap water and 95.27 had BP apparatus. Nine out of ten facilities offering normal vaginal delivery had sterilization equipment examination light (93.81%), resuscitation mask (91.65%) and partograph (90.39). Similarly, two third or more of the facilities had emergency transport (81.49%), sterilization (66.15%), delivery package (74.99%), suction apparatus (65.68%) and infant weighing machine (78.66%). However, fewer than one third of the total facilities had a manual vacuum extractor (23.20%) and vacuum aspirator (20.93%).

Figure 4 illustrates that there is substantial variation in BEmONC service readiness score by district and type of facilities.

**Figure 4:**
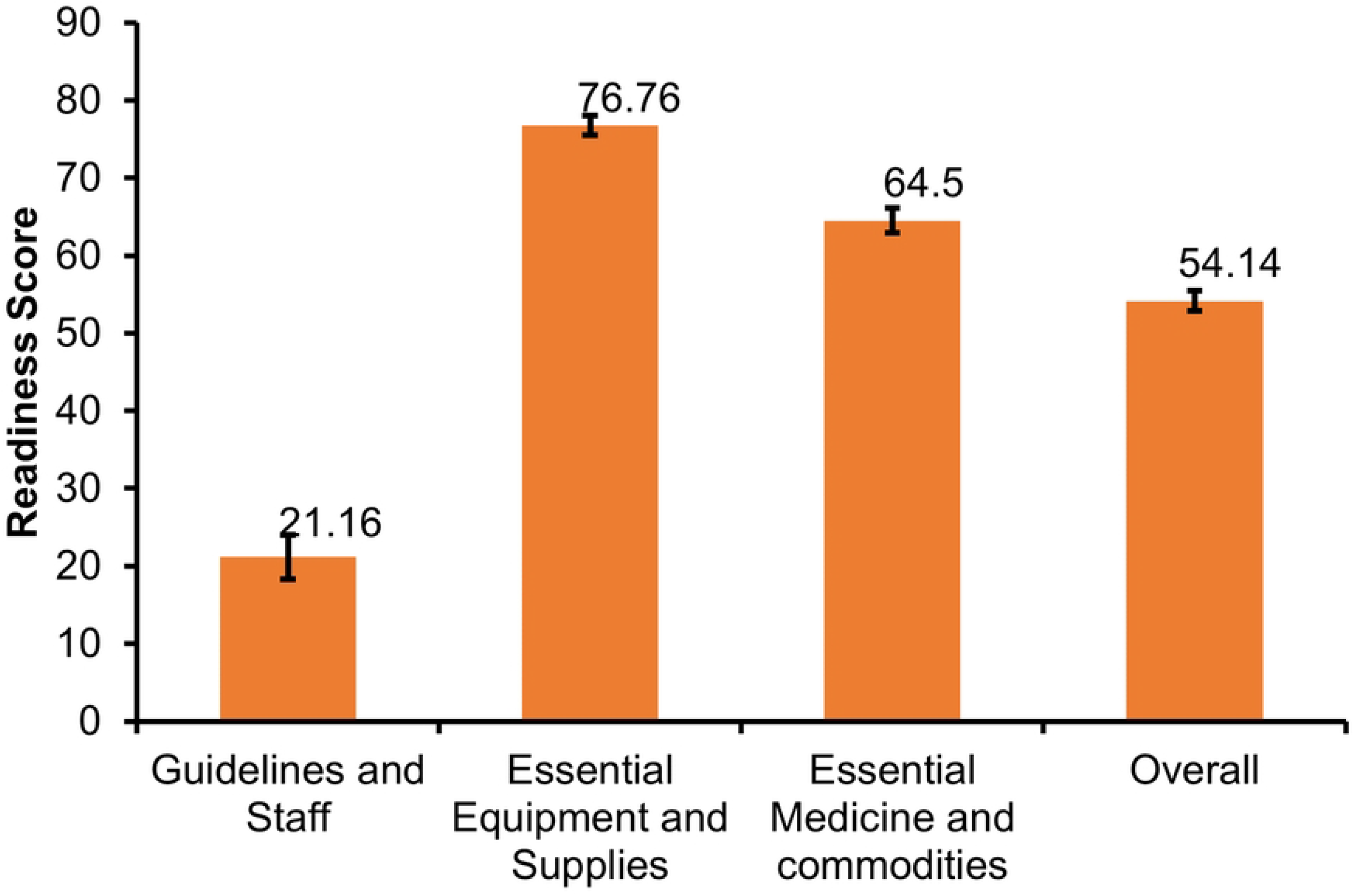
Facility wise and district wise BEmONC readiness.

Gardner-Altman estimation plot showed that readiness score vary marginally by province, ecological region and rural/urban setting. (Figure 5) Factors associated with BEmONC service readiness are presented in Table 6. In bivariate analysis, ecological belt, level of facilities, presence of external supervision, 24-hour duty schedule, quality assurance activities, number of total health workers and delivery beds were significantly associated with BEmONC service readiness score. Similarly, facilities having system for review of maternal and newborn deaths had better readiness score than those not having such review. As per findings from multiple linear regression, with reference to federal/provincial level hospitals, local hospitals/ PHCCs/peripheral facilities (−12.77 points, 95% CI: −18.52, −7.01, p-value: <0.001) and private hospitals have lower readiness score (−18.39 points, 95% CI: −24.475, − 12.308, p-value: <0.001). Compared to Province 1, BEmONC readiness score is 3.49 points higher in Bagmati Province (95% CI: 0.43, 6.55, p-value: 0.03). Similarly, facilities located in rural setting have better service readiness score (2.80 points higher, 95% CI: 0.79, 4.80, p-value: 0.006) than urban facilities and facilities in mountain belt (4.18 points higher, 95% CI: 1.61, 6.74, p-value: 0.001) have better readiness score than facilities in Hilly region. Some of the service management related variables were also associated with better readiness score. For example, facilities having external supervision (3.08 points, 95% CI:1.16, 5.01, p-value: 0.002), and 24 hours duty schedule (5.52 points, 95% CI: 2.99, 8.06, p-value: <0.001). With one unit increase in number of delivery bed, the readiness score increased by approximately one point (0.97 points, 95% CI: 0.08, 1.87, p-value: 0.03).

**Figure 5:**
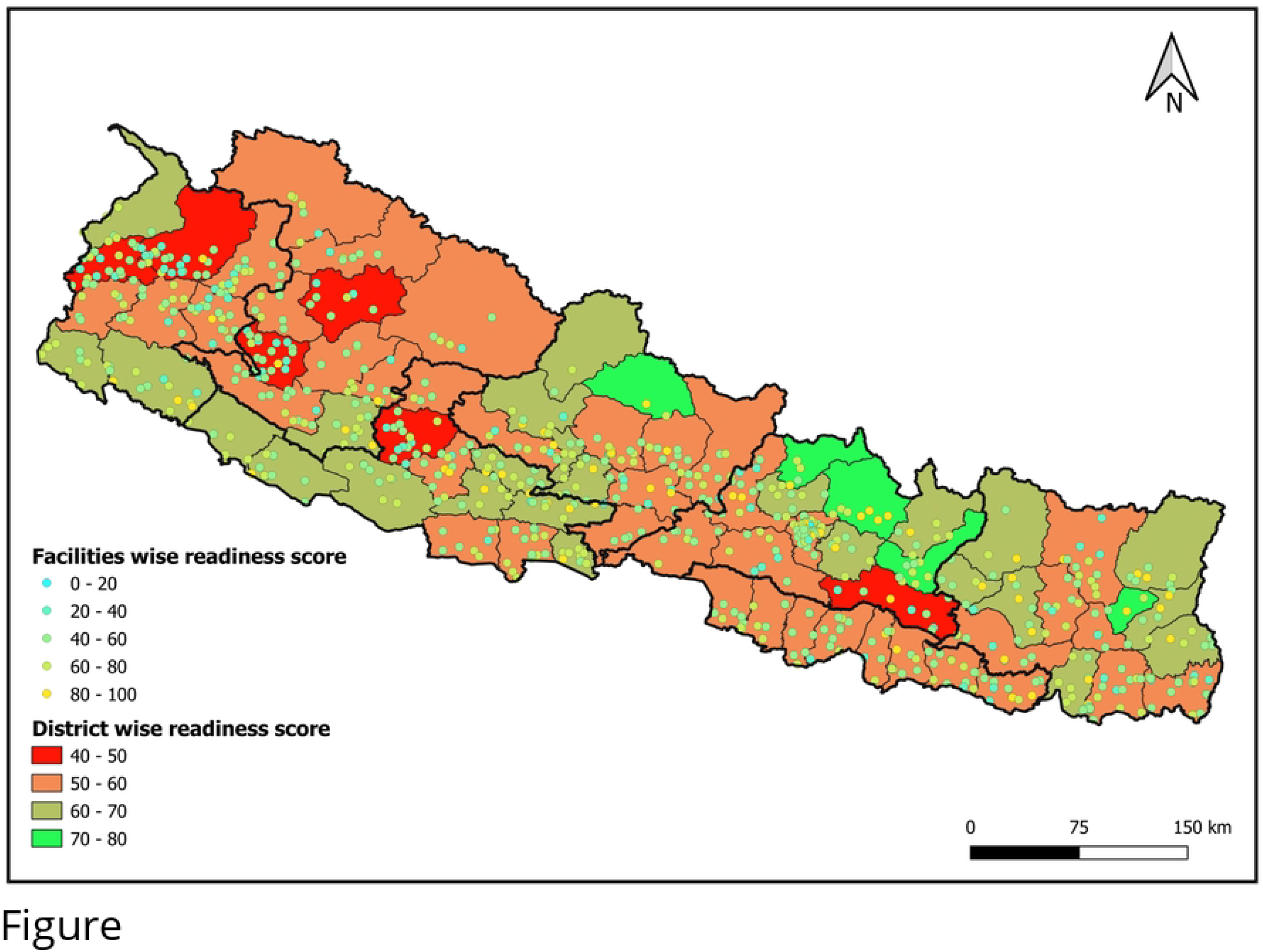
Gardner-Altman estimation plots for comparing readiness score of facilities offering normal vaginal delivery services disaggregated by A) province, B) Ecological region, and C) Rural/Urban setting.

Figure 4 illustrates that there is substantial variation in BEmONC service readiness score by district and type of facilities.

Gardner-Altman estimation plot showed that readiness score vary marginally by province, ecological region and rural/urban setting. (Figure 5)

Factors associated with BEmONC service readiness are presented in Table 6. In bivariate analysis, ecological belt, level of facilities, presence of external supervision, 24-hour duty schedule, quality assurance activities, number of total health workers and delivery beds were significantly associated with BEmONC service readiness score. Similarly, facilities having system for review of maternal and newborn deaths had better readiness score than those not having such review. As per findings from multiple linear regression, with reference to federal/provincial level hospitals, local hospitals/ PHCCs/peripheral facilities (−12.77 points, 95% CI: −18.52, −7.01, p-value: <0.001) and private hospitals have lower readiness score (−18.39 points, 95% CI: −24.475, − 12.308, p-value: <0.001). Compared to Province 1, BEmONC readiness score is 3.49 points higher in Bagmati Province (95% CI: 0.43, 6.55, p-value: 0.03). Similarly, facilities located in rural setting have better service readiness score (2.80 points higher, 95% CI: 0.79, 4.80, p-value: 0.006) than urban facilities and facilities in mountain belt (4.18 points higher, 95% CI: 1.61, 6.74, p-value: 0.001) have better readiness score than facilities in Hilly region. Some of the service management related variables were also associated with better readiness score. For example, facilities having external supervision (3.08 points, 95% CI:1.16, 5.01, p-value: 0.002), and 24 hours duty schedule (5.52 points, 95% CI: 2.99, 8.06, p-value: <0.001). With one unit increase in number of delivery bed, the readiness score increased by approximately one point (0.97 points, 95% CI: 0.08, 1.87, p-value: 0.03).

## Discussion

We evaluated the BEmONC service readiness and availability based on nationwide representative NHFS 2021.

Study findings indicate that for most of the signal functions, availability declined from 2015 to 2021 except the availability of parenteral oxytocin [15]. Among seven signal functions, parenteral oxytocin was available in 88.2% of facilities, which is slightly higher than the availability reported in previous round of survey in 2015 (85.8%) [15]. From 2015 to 2021, the availability of the manual removal of placenta declined to 36.7% from 42.8%, removal of retained product of placenta declined to 26.4% from 33%, the availability of parenteral administration of antibiotics declined to 36.5% from 40.7%, neonatal resuscitation service availability declined to 29.6% from 36.8% and the availability of parenteral anticonvulsants declined to 8.9% from 10%. Notably, the availability of assisted vaginal delivery declined to 8.1% in 2021 from 16.1% in 2015[15]. These findings also tally with the findings presented in Annual report of DoHS. The met need of EOC has been continuously declining in the last four fiscal years. The met need of EOC was 12.6% in 2017/18, 11.6% in 2018/19, 11.1% in 2019/20 and 8.2% in 2020/21 [7].

When compared to other neighboring countries like Pakistan and Bangladesh, Nepal seems to have relatively low availability of BEmONC signal functions. In the Sindh Province study (Pakistan), the proportion of facilities having parenteral antibiotics was 92%, oxytocin was 90%, service for manual removal of placenta was 92%, service for normal birth facilities was 82% and neonatal resuscitation service was 80% [16]. Similarly, 98% of health institutions in Bangladesh had parenteral antibiotics, 90% had oxytocin, 86% had manual placenta removal service, and 92% had health workers who could remove residual products of conception [17].

Although SMNH roadmap has set the target for 100% met need of EOC, the annual report from Department of Health Services reveals that Nepal has relatively poor EOC met need. Between the fiscal year 2017/18 to 2020/21, the nationwide availability of EOC met need dropped from around 11% to roughly 8%. This is a big source of worry, and it may signal a decrease in the use/delivery of complication management throughout pregnancy[7]. In 2020/21, the met need of EOC was 8.2% at national level, 10.2% in Province 1, 3.5% in Madhesh, 10.4% in Bagmati, 4.5% in Gandaki, 10.5% in Lumbini, 6.3% in Karnali and 9.9% in Sudurpaschim Province [7].

In last five years, Nepal witnessed the transition to a federal structure that involved the redefining of the responsibilities of government bodies at federal, provincial and local level. In the current federal structure, there are 753 local governments that assume the responsibility of delivery of basic health services including birthing facilities. Decline in availability of six out of seven signal functions could be due to lack of coordination and clarity of roles between the three tiers of government (1 federal, 7 provincial, and 753 local governments) and limited availability of health workers in health facilities at multiple level because of staff readjustment process in first few years of transition to federal structure. The survey was completed while the COVID-19 pandemic was ongoing. The first case of COVID-19 was seen in Nepal on 23 January 2020. Originally scheduled data collection in 2020 was rescheduled because of the nationwide lockdown imposed to contain COVID-19 pandemic. The restrictive measures imposed to contain the spread of COVID-19 pandemic could have had some impact on the availability of BEmONC signal functions [12]. Similarly, these differences could be partly because of some methodological differences. For example, there is more representation of peripheral and private facilities in the 2021 study compared to the 2015 study.

The aggregate weighted BEmONC service readiness score was 57 and the unweighted service readiness score was 55. Findings show significant difference in BEmONC service readiness based on province, location (urban/rural), ecological belt (mountain, hill and terai), type of facilities (federal/provincial, local), presence of external supervision, 24 hours duty schedule, quality assurance activity and the number of delivery beds. Further analysis from 2015 NHFS also revealed statistically significant variation in service readiness score based on type of health facilities and availability of 24 hour BEmONC services [15]. Facilities in Bagmati Province had higher BEmONC service readiness score (by 3.37 points, p-value=0.034). Unlike previous study [15], the BEmONC service readiness was better in rural area (2.76 points, p-value=0.008) which could be because higher proportion of health workers in rural area have been trained on BEmONC related services [12].

The overall service readiness score in our study was 55, marginally higher than the score (52%) in the previous study [12]. The weighted readiness score for the guideline and staff was 21.3, for equipment was 81.9, and for essential medicine and commodities was 67. The weighted readiness score for guideline and staffs was 28.5, equipment and supplies was 72.2 and essential medicine and commodities was 55.5 [15]. The readiness score for equipment and supplies and the essential medicines and commodities have improved from previous round of survey. The overall readiness score in case of Nepal is higher than that of some other countries like Tanzania and Kenya [18,19].

Among individual domains of readiness score, guideline and staff has the lowest score and thus has highest scope for improvement. A smaller number of training sessions were organized after emergence of COVID-19 pandemic due to restrictive measures limiting gathering of health workers in one place, and this could be one possible reason for the limited number of training sessions organized. Further, redefinition of roles and responsibilities in federal structure and limited understanding among the concerned authorities could be the other reasons. Only one third of facilities had at least one service provider trained on SBA, advanced SBA, maternal and newborn health update, routine care during labor and normal vaginal delivery skills. ATMSL in the last 24 months seems concerning considering the emphasis that has been placed on quality of care in recent years. One of the previous studies indicates that almost one third of the decline in maternal and neonatal mortality (28% decline in each case) could be realized through improvement of the quality of care in low and middle income countries [20].

Findings of our study present a comprehensive understanding of where Nepal stands in terms of service availability and readiness for BEmONC. As the decision-making authority has been devolved to 7 provincial and 753 provincial governments, there could be opportunities for improving readiness, service availability and quality of service in all birthing centres. Further studies that involve in-depth assessment of birthing centres in each province with cost estimation for upgrading health facilities as per the national standards could be useful. Apart from improvement of service readiness and availability, efforts should also be directed to raise awareness at community level to encourage utilization of available health services.

We utilized data from a survey that was designed to assess the service availability and readiness for multiple other conditions like NCDs and was not primarily dedicated to emergency obstetric and newborn care. Thus, study lacks some variables like frequency of pregnancy related complication and outcomes. Furthermore, since the study was undertaken during the period of COVID-19 pandemic, some components of service availability and readiness might have been under-estimated or over-estimated in this study.

## Conclusions

At national level, the proportion of facilities having all seven BEmONC signal functions was very low. The availability of BEmONC signal functions varied notably by province, ecological belt and urban/rural setting, presence of external supervision, 24-hour duty schedule, level of facilities and number of delivery beds. The study findings highlighted the need to enhance the service availability and readiness of BEmONC services and number of service delivery professionals, service hours, and conducting a periodic evaluation of maternal and newborn fatalities.

## Data availability

The dataset is publicly available in the official website of “The Demographic and Health Surveys” program (https://dhsprogram.com/)

## Acknowledgement

We would like to acknowledge United States Agency for International Development’s Demographic Health Survey program for providing us the datasets and thank all who directly and indirectly supported us in this study.

## Competing interest

Authors declare that there is no competing interest

## Notes

### Competing Interest Statement

The authors have declared no competing interest.

### Funding Statement

The author(s) received no specific funding for this work

### Author Declarations

It is further analysis of publicly available Nepal Health Facility Survey 2021 data and no separate ethical clearance is not required

